# Genomic surveillance of SARS-CoV-2 and emergence of XBB.1.16 variant in Rajasthan

**DOI:** 10.1101/2024.01.11.24300881

**Authors:** Pratibha Sharma, Swati Gautam, Abhaya Sharma, Dinesh Parsoya, Farah Deeba, Nita Pal, Ruchi Singh, Himanshu Sharma, Neha Bhomia, Ravi P. Sharma, Varsha Potdar, Bharti Malhotra

## Abstract

**Background & objectives:** Genomic surveillance of positive SARS-CoV-2 samples is important to monitor the genetic changes occurring in virus, this was enhanced after the WHO designation of XBB.1.16 as a variant under monitoring in March 2023. From 5^th^ February till 6^th^ May 2023 all positive SARS-CoV-2 samples were monitored for genetic changes.

**Methods:** A total of 1757 samples having Ct value <25 (for E and ORF gene) from different districts of Rajasthan were processed for Next Generation sequencing (NGS). The FASTA files obtained on sequencing were used for lineage determination using Nextclade and phylogenetic tree construction.

**Results and discussion:** Sequencing and lineage identification was done in 1624 samples. XBB.1.16 was the predominant lineage in 1413(87.0%) cases while rest was other XBB (207, 12.74%) and other lineages (4, 0.2%). Of the 1413 XBB.1.16 cases, 57.47% were males and 42.53% were females. Majority (66.53%) belonged to 19-59 year age. 84.15% of XBB.1.16 cases were infected for the first time. Hospitalization was required in only 2.2% cases and death was reported in 5 (0.35%) patients. Most of the cases were symptomatic and the commonest symptoms were fever, cough and rhinorrhoea. Co-morbidities were present in 414 (29.3%) cases. Enhanced genomic surveillance helped to rapidly identify the spread of XBB variant in Rajasthan. This in turn helped to take control measures to prevent spread of virus and estimate public health risks of the new variant relative to the previously circulating lineages. XBB variant was found to spread rapidly but produced milder disease.

## Introduction

Since the first identification of SARS-CoV-2 outbreak in Wuhan, many variants have emerged due to variations in nucleotides [1]. The recent variant of SARS-CoV-2 generated by the recombination between BJ.1 & BM.1.1.1, named as XBB (Gryphon) belongs to BA.2 lineage [2-5]. In late 2022, XBB.1.5 variant, a descendant lineage of XBB.1, emerged and rapidly spread in U.S.A and other countries [6, 7]. Another XBB sub lineage, XBB.1.16 emerged and was first detected in India in February 2023 and high positivity was observed in end of March 2023. XBB.1.16 was named as Arcturus and was believed to be more transmissible than other variants reported earlier [8]. Genetic profile of XBB.1.16 is similar to XBB.1.5 with two additional amino acid mutations in the spike protein E180V and K478R. The World Health Organization (WHO), on 22^nd^ March 2023 designated XBB.1.16 as a variant under monitoring and later as a variant of interest on 17^th^April 2023 [9]. Looking at the increase in SARS-CoV-2 positivity, the Government of Rajasthan decided to enhance the genomic surveillance to process all positive SARS -CoV-2 samples for Next Generation Sequencing (NGS) from February 2023 onwards.

## Methodology

During the period from 5^th^ February till 6^th^ May 2023, a total of 3460 Nasopharyngeal/Throat swab specimens from SARS-CoV-2 positive patients were received from all over Rajasthan. Demographic and other details of the patients were obtained along with the samples. Out of these, 1757 samples having high viral load (cycle threshold <25 for E and ORF gene) were processed for Next Generation Sequencing (NGS) at a government medical college at Jaipur which is the satellite lab of Indian SARS-CoV-2 Genomics Consortium (INSACOG) and National Institute of Virology (NIV) Pune being the hub lab. Nucleic acid extraction was done on EasyMag extraction system (Biomerieux, Netherlands) and cDNA synthesis was done using Superscript VILO reverse transcriptase kit (Invitrogen, USA). Library preparation was done by Ion AmpliSeq library kit plus, template preparation, enrichment and chip loading was done on Ion Chef system (Life Technologies) and loaded chips were sequenced on the Ion torrent S5 system(Life Technologies). NGS data analysis was done as described previously [10]. The FASTA files obtained on sequencing were aligned withWuhan-Hu-1/2019 (genbank: MN908947) as reference using Nextclade (https://clades.Nextstrain.org). Phylogenetic tree was downloaded and visualised using Nextstrain Auspice (accessed on 24^th^June 2023) web software. The study was approved by the institutional ethics committee.

## Results

Rapid increase in number of XBB.1.16 cases was observed, from epidemiological week (epiweek) 6 (0%) to epiweek18 (90%). The first case of XBB.1.16 was observed in 7^th^epiweekon 14^th^ February’ 2023. A total of 1757 samples were tested during the period from 6^th^epiweek to 18^th^epiweek (5^th^ Feb 2023 till 6^th^ May 2023), out of which 1624 (92.43%) met with quality score and had more than 95% genome coverage. Among the lineages identified, XBB.1.16 was the predominant lineage which was detected in 1413 (87.0%) cases while rest was other XBB (207, 12.74%) and other lineages (4, 0.2%). Among other XBB lineages, XBB.2.3 was 135(8.31%), XBB.1 and XBB.1.9.1 in 14 (0.86%) cases each, XBB.1.5 and XBB.2.3.2 in 11 (0.67%) cases each, XBB.2.3.3 and XBB.1.9.2 in 5(0.31%) cases each, XBB.2.3.5 in 4(0.24%), XBB.2.7.1 in 2(0.12%), and 1 (0.06%) case each of XBB, XBB.1.5.11, XBB.1.5.21, XBB.1.5.24, XBB.2 and XBB.2.3.4.The phylogenetic representation of the lineages identified is given in Figure 1. Maximum positivity for SARS-CoV-2 as per Integrated Disease Surveillance Project (IDSP) data was observed in 15^th^epiweek during which 87.63% were XBB.1.16 cases and 12.36% were other lineages. Out of the total 1413 XBB.1.16 cases, 812 (57.47%) were from males and 601(42.53%) from females. The majority (940, 66.53%) of the XBB.1.16 cases belonged to 19– 59-year age group followed by ≥ 60-year age group (350, 24.77%). Least cases were seen in <12 years age (52, 3.68%). Majority (1275, 90.23%) of the population was vaccinated, 6.09% were unvaccinated while 3.68% were not eligible for vaccination according to their age group (<12 years). Lab confirmed reinfection occurred in 15.85% cases and 84.15% of XBB.1.16 cases were infected for the first time (Table I).

**Table I:** Patient Demographics and Clinical Characteristics of XBB.1.16.

| **Demographic Characteristics (N=1413)** |  |
| --- | --- |
| **Age (Yrs.)** |  |
| <12 | 52 (3.68) |
| ≥12 to 18 | 71 (5.02) |
| 19 to 59 | 940 (66.53) |
| ≥60 | 350(24.77) |
| **Gender** |  |
| Male | 812 (57.47) |
| Female | 601 (42.53) |
| **Area wise Distribution** |  |
| **Ajmer** | 133 (9.41) |
| **Alwar** | 108 (7.65) |
| **Bhilwara** | 28 (1.98) |
| **Bikaner** | 42 (2.97) |
| **Chittorgarh** | 68(4.81) |
| **Churu** | 3(0.21) |
| **Dausa** | 20 (1.42) |
| **Dholpur** | 4(0.29) |
| **Dungarpur** | 13(0.92) |
| **Hanumangarh** | 6(0.43) |
| **Jaipur** | 796(56.33) |
| **Karauli** | 1(0.07) |
| **Nagaur** | 55(3.89) |
| **Rajsamand** | 3(0.21) |
| **Sikar** | 66(4.67) |
| **Udaipur** | 67(4.74) |
| **Clinical characteristics** |  |
| **Lab confirmed reinfection** |  |
| Yes | 224(15.85) |
| No | 1189(84.15) |
| **Symptoms** |  |
| Symptomatic | 1305 (92.35) |
| Non symptomatic | 108 (7.64) |
| **Co-morbid conditions** |  |
| One co-morbidity | 233(16.48) |
| Two co-morbidity | 168(11.88) |
| Three co-morbidity | 13(0.92) |
| **Travel History** |  |
| International | 7 (1.3) |
| National | 14 (2.7) |
| No travel history | 1392 (98.5) |
| **Vaccination History** |  |
| Unvaccinated | 86 (6.09) |
| Vaccinated | 1275 (90.23) |
| Not Eligible | 52 (3.68) |
| ***Clinical Outcome*** |  |
| Hospitalization | 31 (2.2) |
| No Hospitalization | 1382 (97.8) |
| Death | 5(0.35) |
| ***Epiweek wise emergence of XBB.1.16*** |  |
| **Epiweek No.** | **XBB.1.16/Total (%)** |
| 6 | 0/2 (0) |
| 7 | 1/2 (50) |
| 8 | 0/2 (0) |
| 9 | 10/21 (47.62) |
| 10 | 11/12 (91.66) |
| 11 | 60/75 (80) |
| 12 | 62/84(73.81) |
| 13 | 73/84(86.9) |
| 14 | 191/222(86.04) |
| 15 | 411/469 (87.63) |
| 16 | 418/461(90.67) |
| 17 | 158/170(92.94) |
| 18 | 18/20 (90) |

**Figure 1:**
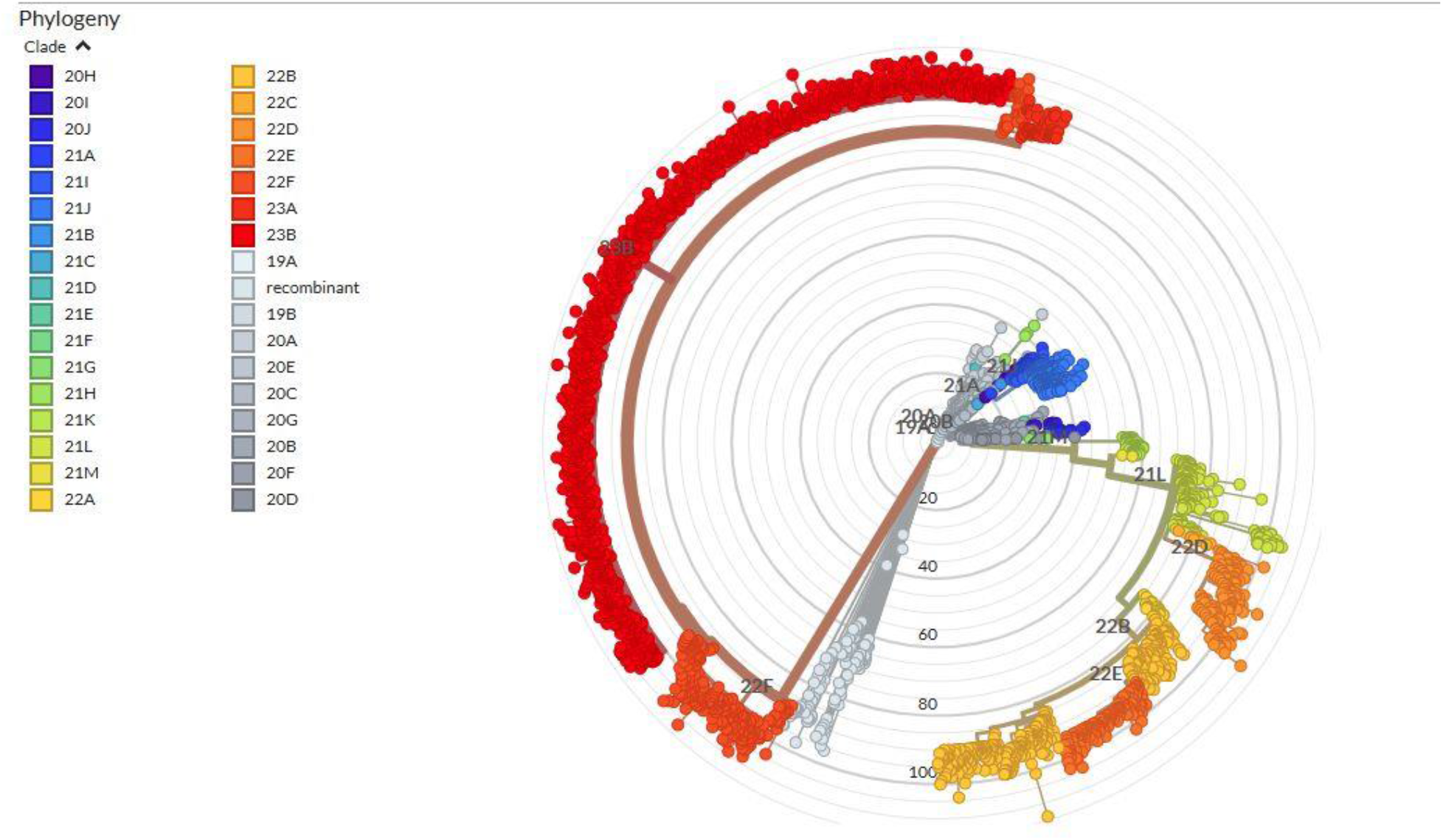
Phylogenetic representation of the SARS-CoV-2 lineages.

Among XBB.1.16 cases, maximum numbers of positive cases were from Jaipur district (796, 56.33%) followed by Ajmer (133, 9.41%), Alwar (108, 7.65%), Chittorgarh (68, 4.81%), Udaipur (67, 4.74%) and Sikar (66, 4.67%) (Table I).

First positive case in Rajasthan was detected on 14^th^ February 2023 in a female patient from Jaipur district with no history of international travel as well as the XBB1.16 prone area within the country. Of the total of 1413 positive cases identified, 7 (1.3%) had a history of international travel among which 4(57.14%) had a history of travel to Australia while 3 (42.86%) to America. Majority (1305, 92.35%) of the cases were symptomatic. The common symptoms observed among those having mild disease were fever (727, 51.45%) and cough (697, 49.33%) followed by rhinorrhea (458, 32.41%), body ache (210, 14.86%), sore throat (121, 8.56%), nausea (37, 2.62%), breathlessness (11, 0.78) and diarrhea (6, 0.42%). Among XBB.1.16 cases, 233 (16.48%) were found to have one co-morbidity, 168 (11.88%) had two co-morbidities and 13 (0.92%) had three co-morbidities. The most common co-morbidities found were diabetes (138, 9.76%), hypertension (108, 7.6%), asthma (155, 10.96 %), thyroid abnormalities (105, 7.4%), heart disease (75, 5.3%), tuberculosis (22, 1.55%), renal disease (2, 0.14%), liver disease (1, 0.07%), HIV (1, 0.07%) and cancer (1, 0.07%). Hospitalization was required in only 2.2% cases having severe disease. Death was reported in 5(0.35%) cases. Among the 5 death cases, the co-morbidities were tuberculosis (1, 20%), cancer (1, 20%), tuberculosis, hypertension & diabetes mellitus (1, 20%), hypertension & diabetes mellitus (1, 20%) and tuberculosis, diabetes mellitus & HIV (1, 20%).

## Discussion

In our study, the first case of XBB.1.16 was observed in 7^th^ epidemiological week of year 2023 (on 14^th^ February) when the total number of SARS-CoV-2 positive cases was low. Sudden increase in SARS-CoV-2 positive cases was observed in short time (IDSP data) which was attributed to XBB.1.16 which had rapidly become the predominant lineage. In Rajasthan maximum positivity was seen in 15^th^epiweek, 87.63% were XBB.1.16 cases and 12.36% were other lineages. A weekly rise in the prevalence of XBB.1.16 was also reported globally from 0.52% during epiweek 9, to16.8% in epiweek 20[11]. As of 5^th^ June 2023, 19847 sequences of the Omicron XBB.1.16 variant were available from 66 countries among which 8086 sequences (40.7%) were from India [12]. This sudden rise in number of XBB.1.16 cases has been attributed to its higher transmission advantage owing to its higher “Effective Reproductive Number” (Re), 1.2 which is 1.17 times higher compared to XBB.1 and XBB.1.5 variants revealing its ability to transmit rapidly as reported earlier [11].

Though SARS-CoV-2 infects people of all ages, we found majority (940, 66.53%) of the XBB.1.16 cases belonged to the 19-59 year age group which might be due to the fact that the population in this age group is more active and interacts more socially and in work place leading to higher risk of exposure to infected environments. However, a study from Maharashtra reported 20–39 year age group to be the predominantly affected age group [13].In our study, least cases were observed in <12 years age (52, 3.68%) despite the fact that the population in this age group was not vaccinated due to their non eligibility for vaccination. Lab confirmed reinfection was observed in only 15.85% cases despite the fact that all of them were vaccinated. This might be an underestimate of the actual reinfection cases which were not reported owing to the asymptomatic clinical presentation. These findings are similar to those reported from Maharasthra where the reinfection among XBB.1.16 cases was found to be 10.86% [13]. XBB.1.16 is reported to be more immune evasive when compared to other XBB variants like XBB.1 and XBB.1.5 [14] and this might be one of the reasons for re-infection among the vaccinated individuals as observed in our study. Most of the infected cases in our study were symptomatic and the common symptoms observed were fever, cough and rhinorrhea which are similar to those reported in our previous study on Omicron [10].Study by Vashishtha *et al* reported fever (100%), rhinorrhea (72.3%) and cough (59.1%) to be the most common symptoms among pediatric age group [15]. Study from Maharashtra reported 92% of the XBB.1.16 infected cases to be symptomatic with fever (67%) being the most common symptom [13]. Cobar *et al* in their report mentioned high fever, sore throat, loss of senses of smell and taste, headache, muscle pain accompanied by cramps, fatigue and conjunctivitis to be the common features presented in individuals infected with XBB.1.16 variant [11].

Co-morbidities were present in 29.3% cases in our study. These findings are similar to those from Maharashtra where co-morbidities were reported in 17.7% of cases, of which hypertension was the most common (47.9%), followed by diabetes mellitus (39.6%) and asthma (12.5%) [13].Study from Chennai, Tamil Nadu reported diabetes (38%), hypertension (28%) and cardiovascular disease (13%) to be the significant co-morbidities present in patients infected with XBB variant [16]. Among XBB.1.16 cases, maximum number of positive cases were from Jaipur district (796/1413, 56.33%) followed by other districts which might be attributed to high positivity in Jaipur and timely transport of positive samples to sequencing laboratory.

Hospitalization due to COVID disease was required in only 2.2% cases and death was reported in 5(0.35%) cases in our study. IDSP Jaipur reported a total 60 deaths, case fatality rate was 0.59% all over Rajasthan during this period due to COVID. The recovery rate reported for XBB.1.16 variant was 98.75% by Ministry’s of India [17]. Till now, there is no evidence which suggests that XBB.1.16 variant is more severe as no noticeable increase in hospitalizations, ICU admissions, or deaths have been reported due to this variant [18]. Study from Maharashtra also reported no significant difference in hospitalization and disease severity for XBB.1.16 as compared to other co-circulating variants [13]. However, it is essential to remain cautious and to strictly follow preventive measures owing to high transmissibility rate of this virus. It is also important to carry out regular genomic surveillance to monitor for the genetic changes occurring in the virus which will help to estimate public health risks relative to the other currently co-circulating Omicron descendent lineages. Genomic surveillance will be an evidence based approach to contain rapid spread of the virus before its rapid spread causing pandemic situation.

## Data Availability

All data produced in the present study are available upon reasonable request to the authors

